# Saliva cell-free mitochondrial DNA (cf-mtDNA) response during physical and cognitive stress

**DOI:** 10.64898/2026.07.02.26356953

**Authors:** David Shire, Tian Wang, Shuang Wang, Temmie Yu, Martin Picard, Annie T. Ginty, Caroline Trumpff

**Affiliations:** Department of Psychiatry, Division of Behavioral Medicine, Columbia University Irving Medical Center, New York, NY 10032 USA; Department of Biostatistics, Columbia University Mailman School of Public Health, New York, NY 10032 USA; Department of Neurology, H. Houston Merritt Center, Columbia Translational Neuroscience Initiative, Columbia University Irving Medical Center, New York, NY 10032 USA; Robert N. Butler Columbia Aging Center, Mailman School of Public Health, Columbia University, New York, NY 10032 USA; Department of Psychology and Neuroscience, Baylor University, Waco, TX 76798 USA

## Abstract

Emerging evidence suggests that saliva cell-free mitochondrial DNA (cf-mtDNA) increases in response to psychosocial and physical stress. Here, we quantified saliva cf-mtDNA changes in response to acute physical and cognitive stressors as well as identifying potential predictors of these responses, while also exploring the potential modulatory effects of transcranial infrared laser stimulation (TILS). In a crossover design, a total of 47 participants (53% female, ages 18-30) underwent up to three experimental sessions, including an exercise stress task and two cognitive stress tasks. Repeated saliva samples were collected for cf-mtDNA and cell-free nuclear DNA (cf-nDNA) quantification, alongside continuous measurement of heart rate, oxygen consumption, and blood pressure. Our results show that average cf-mtDNA levels increased by 90% after baseline during exercise experiments, and in cognitive stress experiments peaked 160% above average baseline levels during the stress task. Inter-individual differences in response trajectories were associated with differences in factors such as fitness, sleep quality, and stress perception. Notably, participants with higher cf-mtDNA elevations during the exercise experiment reported fewer recent stressful incidents, drank alcohol less frequently, had higher maximum VO_2_ during exercise, and had lower BMI. More dynamic responses to cognitive stress were observed in participants with poorer sleep quality and greater blood pressure reactivity. These findings provide a foundation for larger studies by highlighting the dynamic behavior of saliva cf-mtDNA following physical and cognitive stressors, and by suggesting potential drivers of individual differences in saliva cf-mtDNA stress reactivity.

## 1. Introduction

Acute stress elicits a cascade of physiological and molecular responses aimed at maintaining homeostasis^1^. While short-term stress responses are adaptive, chronic activation of the stress response can contribute to physiological dysregulation and disease risk^2^. However, the molecular pathways linking acute stress adaptation to long-term health outcomes remain incompletely understood^3^.

Mitochondria play a central role in these processes, transforming energy to meet the metabolic demands of stress adaptation^4^. Beyond their intracellular functions, emerging evidence suggests that whole mitochondria can be released into the circulation, where they may contribute to resilience and tissue repair^5,6^, supporting allostasis and energy-intensive adaptive responses^7^.

To measure this phenomenon, cell-free mitochondrial DNA (cf-mtDNA) can be quantified in biofluids. Cf-mtDNA represents fragments or entire mitochondrial genomes that exist in “free” form in the circulation, outside of cells but packaged in different membranous structures, including extracellular vesicles and whole mitochondria^5,8-12^.

In line with the hypothesis that whole mitochondria can be released in the circulation in response to a challenge, several studies have found that acute exercise increases blood cf-mtDNA^13,14^ (reviewed in ^12^). Several studies have also found that acute psychological stress induced an increase in serum and plasma cf-mtDNA levels after 5-45 minutes^15-17^. However, a recent randomized crossover experiment on 72 men and women found that serum cf-mtDNA increased equally during sessions either with or without a socio-evaluative stressor, despite clear mood, cardiovascular, and neuroendocrine responses that were exclusive to sessions with the stressor. This finding calls into question whether prior observations of blood cf-mtDNA levels increasing after socio-evaluative stress may be attributed to procedural factors (such as IV catheter placement or social isolation during the recovery period) rather than acute psychological stress per se^18^.

Saliva provides a non-invasive alternative for measuring cf-mtDNA^19^, avoiding potential confounding effects associated with venipuncture. Two prior studies suggest that acute psychological stress can increase saliva cf-mtDNA levels^17,20^. In a study of 68 women and men participants, acute psychological stress exposure led to an average 280% increase in cf-mtDNA^20^. In a smaller study of 42 women and men participants, exposure to a modified Trier social stress test (TSST) was associated with a 50% increase (non-significant trend) in saliva cf-mtDNA^17^. The authors of the latter study also investigated the response of cf-mtDNA to brief exhaustive exercise stress and found no significant change over time, a finding which remains to be confirmed by other studies. Furthermore, while saliva cf-mtDNA might offer a non-invasive and accessible way of assessing psychological stress responses, its regulation and relationship to other physiological markers of stress, such as heart rate, oxygen consumption, and blood pressure, have not been comprehensively characterized.

In this study, we investigated saliva cf-mtDNA responses to acute physical and cognitive stress challenges. Additionally, we explored the potential buffering effects of transcranial infrared laser stimulation (TILS), a novel neuromodulation technique^21,22^, on cf-mtDNA responses during cognitive stress. We also examined whether saliva cf-mtDNA responses to exercise and cognitive stress challenges were influenced by demographic, behavioral, and physiological factors. To do so, we used advanced statistical approaches, including latent class linear mixed models (LCLMM) to define distinct cf-mtDNA response trajectories and identify their key determinants.

## 2. Methods

### 2.1. Participants and Procedures

Participants were recruited from the greater Waco, Texas area (n=47, 53% female, ages 18-30 years). Exclusion criteria were history of chronic medical or neurological disorder, current illness or infection, pregnancy, and any condition that would prohibit physical exercise. Participant demographics are summarized in Table 1. Participants underwent up to three experimental sessions. The first of these was an exercise stress task, where participants pedaled a stationary bike over several periods of increasing resistance. In the second and third sessions, participants were conditioned with either TILS or a ‘sham’ treatment where they were not exposed to laser light, before performing a task designed to elicit cognitive stress. All experimental sessions began with a 10-minute baseline period and ended with a 30-minute recovery period. Participants were instructed to abstain from vigorous exercise and alcohol for 12 hours and caffeine, food, and drink (except water) for 2 hours prior to the visit.

**Table 1.** Participant demographics separated by experiment.

|  | **Exercise experiment** | **Initial cognitive stress experiment** | **Follow-up cognitive stress experiment** |
| --- | --- | --- | --- |
| **Number of Participants** | 47 | 40 | 39 |
| **Sex, No. (%)** |  |  |  |
| **Female** | 25 (53%) | 20 (50%) | 19 (49%) |
| **Male** | 22 (47%) | 20 (50%) | 20 (51%) |
| **Age (years)\*** | 21±2.2 | 21±2.3 | 21±2.4 |
| **Race, No. (%)** |  |  |  |
| **American Indian or Alaskan Native** | 2 (4.2%) | 2 (5%) | 2 (5%) |
| **Asian** | 11 (23.4%) | 9 (22.5%) | 9 (23%) |
| **Black or African American** | 4 (8.5%) | 4 (10%) | 4 (10.2%) |
| **Native Hawaiian or Other Pacific Islander** | 0 | 0 | 0 |
| **White** | 23 (48.9%) | 20 (50%) | 19 (48.7%) |
| **Mixed** | 7 (14.9%) | 5 (12.5%) | 5 (12.8%) |
| **Ethnicity, No. (%)** |  |  |  |
| **Not Hispanic or Latino** | 36 (76.6%) | 31 (77.5%) | 30 (77%) |
| **Hispanic or Latino** | 11 (23.4%) | 9 (22.5%) | 9 (23%) |
| **Body Mass Index\*** | 24.1±3.8 | 24.3±3.9 | 24.2±4.0 |

\* Data shown as mean (standard deviation)

In total, 47 participants completed the exercise stress experiment, 40 completed an initial cognitive stress experiment, and 39 completed a follow-up cognitive stress experiment. On average, participants completed the first cognitive stress experiment 8.95±5.31 days after the exercise experiment and the follow-up cognitive stress experiment 7.13±5.71 days after the first.

#### 2.1.1. Exercise Stress Experiment Protocol

Participants’ heights, weights, and waist and hip circumferences were measured. Participants were instructed to sit quietly and relax for 5 minutes to acclimate to the environment. They were then fitted with electrodes to measure heart rate (HR) (MindWare Mobile Impedance Cardiograph, Model 50-2303-00) and a face mask to measure oxygen consumption (VO_2_) and collect saliva (Parvo Medics TrueMax 2400). Pulse wave velocity was also measured at this time. Participants were instructed to relax and acclimate to the equipment for 10 minutes. This was followed by a 10-minute baseline period, at the start of which HR and VO_2_ measurements and saliva collection began.

Participants then exercised on a stationary bike (Monark, Sweden) for four 4-minute periods of increasing resistance (15W, 30W, 60W, and 90W). Participants were instructed to maintain a pace of 50 revolutions per minute. After these four periods, a ‘maximal’ exercise portion began, in which resistance increased by 30W every minute until participants decided they could no longer maintain pedaling tempo. Participants were then allowed to cool down on the bike for 1 minute without resistance and drink half a cup of water if they chose to. This was followed by a recovery period during which participants sat still for 30 minutes while completing questionnaires about demographics, personality, mood, and behavior (including the Pittsburgh Sleep Quality Index^23^, the Hospital Anxiety and Depression Scale^24^, the Perceived Stress Scale^25^, the Emotional Regulation Questionnaire^26^, Adverse Childhood Experiences Questionnaire^27^, and the Undergraduate Stress Questionnaire^28^). Saliva was collected at a maximum of 15 time points: 0, 5, and 10 minutes from the start of the baseline period, at 0, 4, 8, 12, and 16 minutes from the start of the exercise period (coincident with the start of each intensity level and the end of the 90W period), and at 0, 5, 10, 15, 20, 25, and 30 minutes from the start of the recovery period. HR and VO_2_ were measured continuously.

#### 2.1.2 Cognitive Stress Experiment Protocol

During the second and third visits, participants performed a task designed to elicit cognitive stress after preconditioning with TILS or sham treatment. For these visits, participants were fitted with the same instruments as in the exercise stress visit as well as an automatic sphygmomanometer (Carescape, V100, General Electric, El Paso, Texas, USA) which measured blood pressure (systolic blood pressure SBP; diastolic blood pressure DBP; mean arterial pressure MAP) every two minutes. Participants’ heights, weights, and circumferences at waist and hip were again measured. The acclimation and baseline periods were the same as in the exercise visit. After the baseline period, participants were instructed to sit still for conditioning with TILS (or sham treatment).

In the TILS condition, an infrared laser was applied to the right side of the forehead to stimulate cytochrome oxidase activity in the dorsolateral prefrontal cortex^22^. The laser operated at a wavelength of 1064 nm, with a power density of 250 mW/cm² and an energy density of 120 J/cm² (CG-5000 laser, Dallas, Texas). In the ‘sham’ condition, laser emission was blocked by a shutter in a manner that was indistinguishable to participants. The order of TILS and sham conditions was randomized to avoid order effects.

After 8 minutes of conditioning, participants performed the multi-source interference task (MSIT), which was designed to elicit cognitive stress and has been demonstrated to activate control and attention regions of the brain^29^. The MSIT has been used in acute psychological stress paradigms and reliably evokes cardiovascular stress responses^30-33^. Before starting, participants were told that they were being scored against other participants, and that it was important for them to perform at their best ability. They were told that their scores would be displayed openly so that they could be compared to others, and that they were being videotaped by a conspicuous video camera so that their body language could be interpreted to measure their anxiety. The MSIT consisted of eight 60-second phases which alternated between easy and hard conditions. Participants were instructed to press buttons marked “1”, “2”, “3”, or “4” to indicate which number among four shown on a screen was different from the other three. For example, if the screen showed “1 2 2 2”, the participants would press button “1”. During the congruent phase of the task, the order of the digits was consistent with the order of the buttons. During the incongruent phase, the order of the digits was inconsistent with the order of the buttons. For example, the pattern “1 3 1 1” requires participants to press button “3”, although the second displayed digit is the one which is different from the other two. The MSIT was followed by a 30-minute recovery period, as in the exercise experiment. Saliva was collected at a maximum of 16 time points: 0, 5, and 10 minutes from the start of the baseline period, at 0, 4, and 8 minutes from the start of the conditioning period, at 0, 5, and 9 minutes from the start of the MSIT, and at 0, 5, 10, 15, 20, 25, 30 minutes from the start of the recovery period. HR and VO_2_ were measured continuously, while blood pressure was measured every two minutes during baseline and every minute during the MSIT. Blood pressure was not recorded during the TILS/sham conditioning phase.

### 2.2. Saliva collection and processing

Saliva was collected via passive drool into a tube integrated into the face mask used to measure VO_2_. Saliva samples were centrifuged at 1,000 xg for 5 minutes. Saliva supernatants were transferred to clean tubes and centrifuged at 5,000 xg for 10 minutes. The resulting 5,000 xg supernatants were aliquoted and stored at -80°C before mailing to Columbia University Irving Medical Center for further processing and analysis. Saliva from experiments with fewer than 10 samples (66.6% of time points in exercise experiments, 62.5% of time points in cognitive stress experiments) was not sent for analysis.

### 2.3. cf-mtDNA and cf-nDNA detection

Mitochondrial and nuclear DNA in cell-free saliva were quantified using the MitoQuicLy method^34^. Briefly, aliquots of cell-free saliva were diluted 20x into a lysis buffer (114 mM Tris-HCl; 10% Tween-20; 0.2 mg/ml Proteinase K, Thermofisher #AM2548) and thermolysed overnight (∼16 hours at 55°C followed by 10 minutes at 95°C) in duplicate. All samples from an individual participant were processed and analyzed together to mitigate batch effects. qPCR reactions for each replicate lysate sample consisted of 8 µL lysate, 10 µL TaqMan Fast MasterMix (LifeTech #4444965) and 2 µL of primers and probes (details in Supplemental Table 1) targeting mitochondrial gene ND1 (NADH dehydrogenase subunit 1) and nuclear gene B2M (beta-2-microglobulin). Reactions were run in triplicate on 384-well plates. An eight-step 1:4 serial dilution series of DNA extracted from human fibroblast cultures was also added to each 384-well plate in triplicate as a standard curve; this standard had previously been quantified using digital droplet qPCR, thus permitting absolute quantification of qPCR target genes. Reactions were run and measured using a QuantStudio 7 Flex system (Applied Biosystems; cycling conditions 50 °C for 2 min; 95 °C for 20 seconds; followed by 40 cycles of 95 °C for 1 second and 60 °C for 20 seconds; C_T_ threshold 0.08). The average of each triplet of cycle threshold (C_T_) measurements was used for quantification. When the coefficient of variance (CV) of a triplet was >0.5%, the most outlying value was omitted and the average of the remaining two values was used for quantification. Average C_T_ values were converted to gene quantities (natural log-transformed (LN) copies / µL) by subtracting the intercept and dividing by the slope of a linear regression of the standard curve on the corresponding 384-well plate. The averages of these measurements from pairs of replicate plates were used in subsequent quantitative and statistical analyses. Samples were re-run starting from the lysis stage if the CV of their measurements between replicates was >4.0%. If a sample’s CV between replicates remained >4.0% after re-analysis, the measurement with the lower CV was used in subsequent analyses. Cf-mtDNA was quantified in at least one saliva sample for 37 exercise experiments, 37 initial cognitive stress experiments, and 35 follow-up cognitive stress experiments.

### 2.4. Data processing and statistical analysis

To match the number of saliva collection time points, physiological measurements recorded over several minutes were averaged as described in Table S2. Prior to statistical analysis, ‘reactivity’ values for each visit were calculated from physiological measurements (SBP, DBP, MAP, VO_2_, HR, RMSSD). Reactivity for a particular experiment was defined as the difference between a participant’s maximum post-baseline value (time points 4 through 15 in the exercise experiment, and time points 4 through 16 in the cognitive stress experiments) and the average of that participant’s available baseline values. Because cf-DNA measurements were found to have a lognormal distribution (based on D’Agostino & Pearson tests of measurements at each time point in exercise and cognitive stress experiments; normal-versus-lognormal likelihood test results of ≤5.42E-11; GraphPad Prism version 10.4.2^35^), log-transformed cf-DNA values were used in statistical tests.

To evaluate temporal changes in cf-DNA and physiological measurements during the exercise experiment, we fitted mixed-effects models in GraphPad Prism, with cf-DNA or the physiological measurement as the outcome, time as a fixed effect, and participant as a random effect. To assess whether temporal changes differed between treatment conditions in the cognitive stress experiments, we similarly fitted a mixed-effects model including time as a fixed effect, treatment group indicator (TILS vs. sham), and a time-by-treatment interaction term, with participant as a random effect. For both the exercise and cognitive stress experiments, we performed post hoc testing to assess whether cf-DNA and physiological measurements differed significantly between baseline and any subsequent time points. Dunnett’s test was used to control the family-wise error rate for multiple comparisons against the baseline time point (Dunnett’s p).

To identify potential subgroups with similar cf-mtDNA trajectory patterns and investigate factors associated with individual cf-mtDNA responses to the experimental protocols, participants from the exercise experiment and the initial and follow-up cognitive stress experiments (with TILS and sham groups combined) were separately clustered based on their cf-mtDNA trajectories using latent class linear mixed models (LCLMM). Models were fitted using the ‘hlme’ function from the package ‘lcmm’ (version 2.1.0) in R (version 4.3.1). In the LCLMM, log-transformed cf-mtDNA values were used as the outcome and were normalized by dividing by the within-trajectory mean before analysis. Time was modeled as a nonlinear effect using a third-degree polynomial, and participant was included as a random effect. We assumed that the trajectories could be separated into two to five latent classes and selected the two-class model for further analysis based on the Bayesian information criterion (BIC; Table S3). Trajectories where cf-mtDNA was quantified in fewer than 10 samples (66.6% or 62.5% percent of the maximum possible number of samples for exercise and cognitive stress experiment, respectively) were omitted from this analysis (14% of exercise trajectories, 5% of initial cognitive stress trajectories, and 6% of follow-up cognitive stress trajectories).

We compared subpopulations in terms of their demographics, physiological reactivities, biometric measurements, and questionnaire responses. Comparisons were made in R using t-tests (to compare the means of continuous variables), Fisher’s exact test (to compare the significance of 2x2 contingency tables), and Armitage trend test (to compare the significance of categorical data with >2 outcomes). Hedge’s *g* was calculated to estimate the effect sizes of continuous variables. The effect sizes of categorical variables with binary and multiple outcomes were estimated by *phi* coefficient and Cohen’s *w*, respectively, using the corresponding functions from the ‘effectsize’ package (version 1.0.0) in R.

## 3. Results

### 3.1. Physiological and saliva cf-DNA responses to exercise

We evaluated whether salivary cf-DNA and physiological measures changed over the course of the exercise experiment. Physiological responses (HR and VO_2_) and saliva cell-free DNA dynamics were measured over a time course that included a 10-minute baseline period, 16 minutes of exercise on a stationary bike with increasing intensity, and a 30-minute recovery period in 47 participants (Figure 1A). As expected, physiological measurements showed robust responses to increasing physical exertion throughout the exercise protocol. HR and VO_2_ exhibited significant temporal changes (p<0.0001) (Figure 1B,C) and reached peak values at the conclusion of the highest resistance period. On average, HR slowly recovered by >80% over the 30-minute recovery period, while VO_2_ returned to near-baseline levels within 15 minutes. Both HR and VO_2_ were significantly greater than baseline (time 0) from the start of the exercise phase through the end of the experiment (Dunnett’s p from 0.03 to <0.0001).

**Figure 1.**
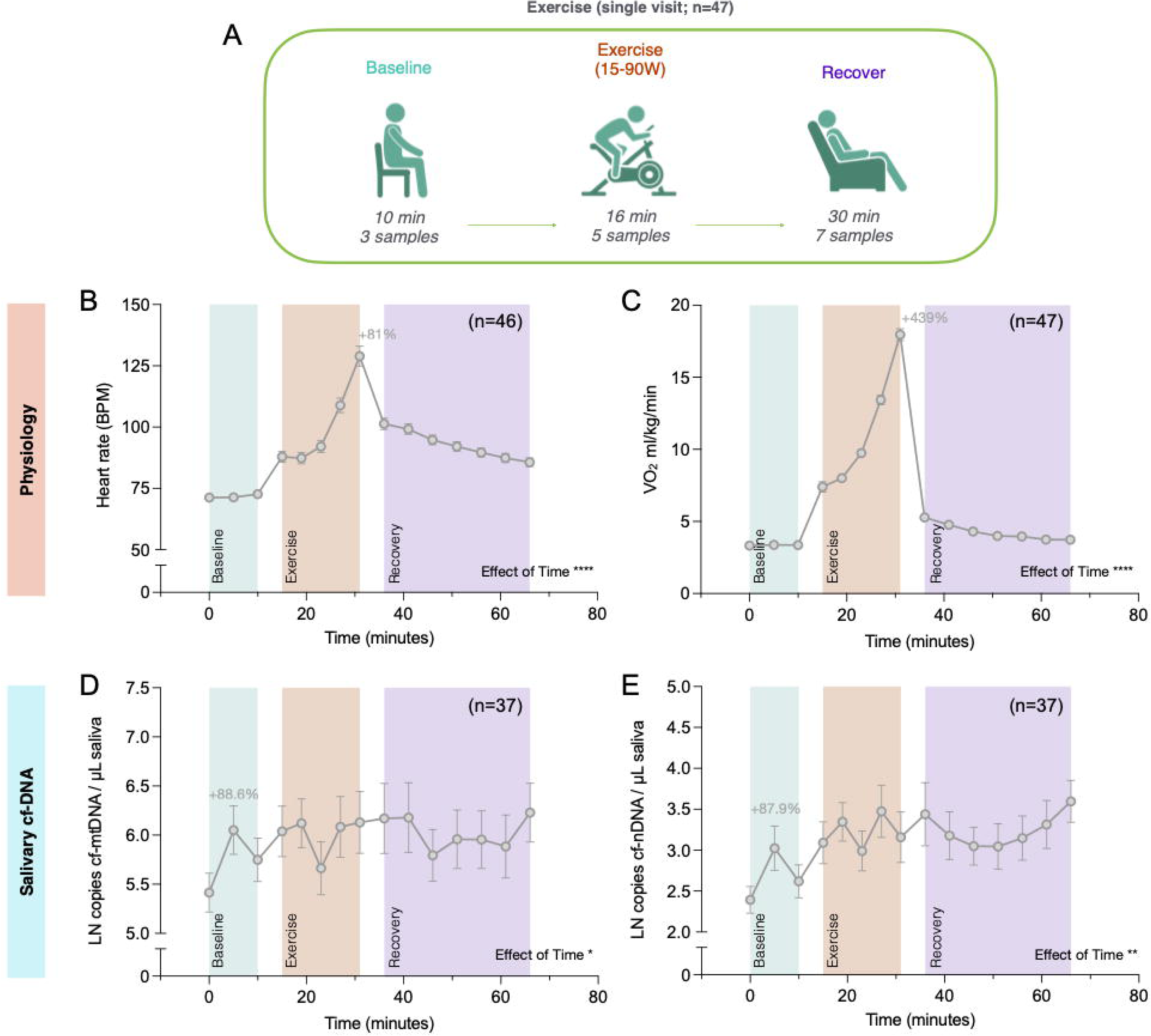
Physiological and saliva cf-DNA reactivity to exercise stress. **(A)** Overview of the experimental design. After a 10-minute baseline period, participants exercised on a stationary bike over four periods of increasing resistance, followed by a 30-minute recovery period. Saliva was collected for cf-mtDNA analysis by passive drool at 15 time points. Heart rate (HR) and oxygen consumption (VO_2_ ml/kg/min) were measured continuously. Trajectories of **(B)** HR and **(C)** VO_2_ ml/kg/min over the exercise stress experiment. Values represent averages of several measurements recorded between indicated time points (details are provided in Table S2). Percentages indicate difference between baseline value (t=0) and value at final exercise time point (t=31). Trajectories of salivary **(D)** cf-mtDNA and **(E)** cf-nDNA abundance over exercise visits. Percentages indicate difference between t=0 and t=5. Values are presented as averages of log-transformed abundance. (B-E) Data shown as average ± standard error of the mean (SEM). Effect sizes and p-values from one-way ANOVA. *p<0.05, **p<0.01, ***p<0.001, ****p<0.0001.

Saliva cell-free mitochondrial DNA (cf-mtDNA) showed significant temporal changes throughout the exercise experiment (p=0.014; Figure 1D). On average, cf-mtDNA showed an 88.6% elevation following the initial baseline measurement, between timepoints one and two. This level was then sustained during the exercise and recovery phases of the experiment. Cf-mtDNA was significantly greater than at 0 minutes at all time points except for 36 and 51 minutes (Dunnett’s p from 0.035 to 0.0007). In contrast to physiological measurements, cf-mtDNA levels did not show reactivity or a dose-response to exercise intensity.

Saliva cell-free nuclear DNA (cf-nDNA) concentrations were 20 to 30-fold lower than cf-mtDNA. However, the relative magnitude of temporal variation of cf-nDNA (p=0.0054; Figure 1E) was similar to that of cf-mtDNA. Cf-nDNA initially increased 87.9% post-baseline and remained elevated throughout the exercise and recovery phases. As with cf-mtDNA, cf-nDNA levels did not increase with exercise intensity.

Normalizing physiological and cf-DNA values to corresponding baseline values (percent change from time 0) did not result in substantially different trajectories or statistical results (Figure S1).

We examined the ratio of cf-mtDNA/cf-nDNA to estimate the portion of cf-mtDNA independently released from cf-nDNA. We found that the cf-mtDNA/cf-nDNA ratio changed significantly over the course of the exercise experiment (p=0.0057), exhibiting a general downward trend from baseline to the end of the recovery period (Figure S2A). When measurements were normalized to baseline values, the average cf-mtDNA/cf-nDNA ratio trajectory was similar to that of non-normalized values, but the change over time was not statistically significant (p=0.08, Figure S2B).

### 3.2. Physiological and saliva cf-DNA responses to cognitive stress

Physiological responses (HR, VO_2_, blood pressure, and heart rate variability) and saliva cell-free DNA dynamics in response to cognitive stress were measured in 40 participants over a time course that included a 10 minute baseline period, an 8 minute period where participants were conditioned with true or sham TILS, a 9 minute computer-based task designed to elicit cognitive stress, and a 30 minute recovery period (Figure 2A). 39 participants also completed a follow-up visit with the same experimental design but with the opposite conditioning treatment from their initial visit (sham or TILS).

**Figure 2.**
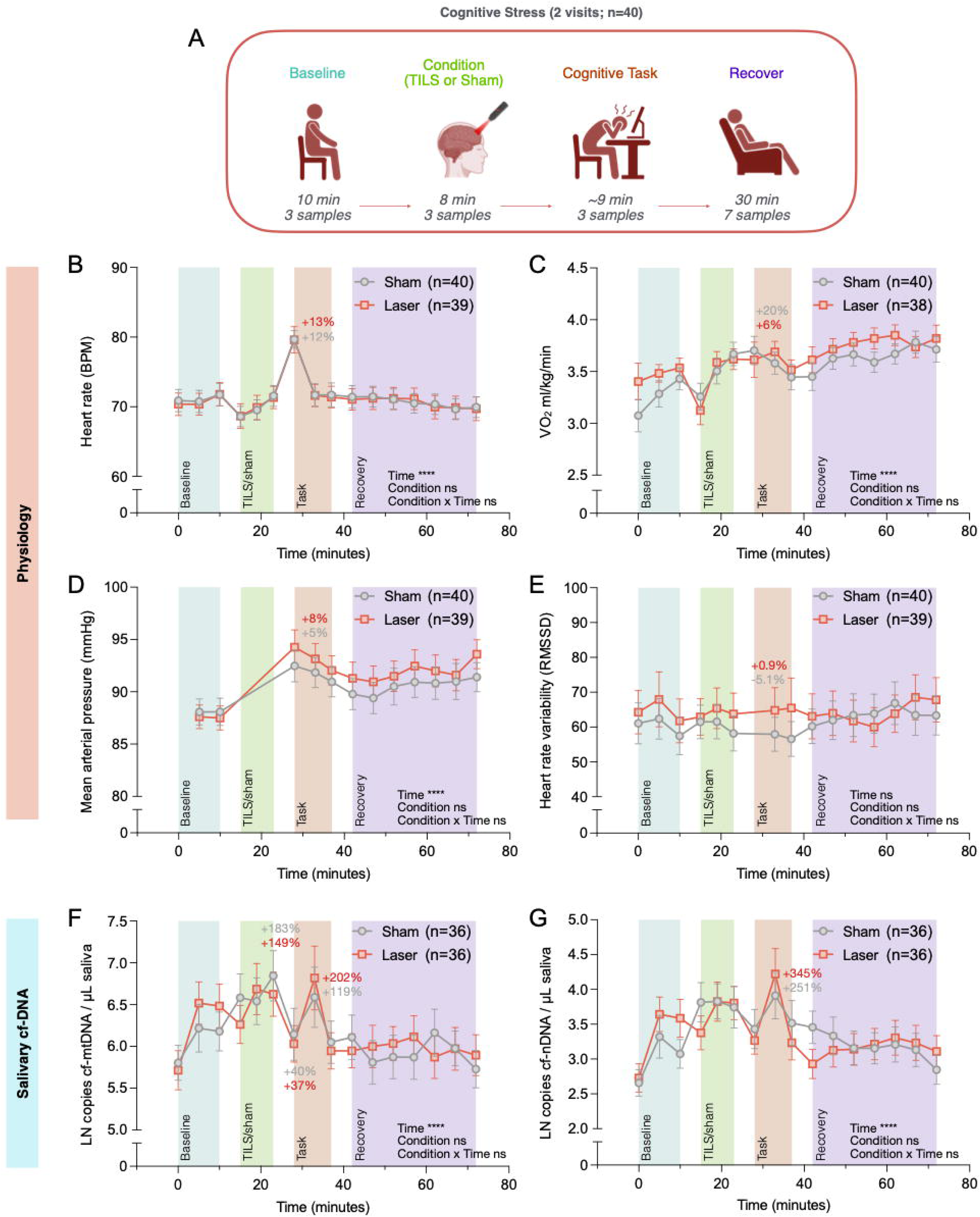
Physiological and saliva cf-DNA reactivity to cognitive stress. **(A)** Overview of the experimental design. After a 10-minute baseline period, participants were exposed to either a transcranial infrared laser stimulation (TILS) or to a sham laser to the right side of the forehead (with the sham/laser visit order randomized) before performing a cognitive task. This was followed by a 30-minute recovery period. Saliva was collected by passive drool for cf-mtDNA analysis at 16 time points. Heart rate (HR) and oxygen consumption (VO_2_ ml/kg/min) were measured continuously. Blood pressure was measured continuously except during the TILS/sham conditioning period. Trajectories of **(B)** HR, **(C)** VO_2_ ml/kg/min, **(D)** mean arterial pressure, and **(E)** heart rate variability (as root mean square of successive differences; RMSSD) over the cognitive stress experiments. Values represent averages of several measurements recorded between indicated time points (details are provided in Table S2). Percentages indicate difference between baseline value (t=0) and value at first task time point (t=28). Salivary **(F)** cf-mtDNA and **(G)** cf-nDNA trajectories over the cognitive stress experiments. Percentages indicate difference between geometric mean of baseline value (t=0) and values at second task time point (t=33). (B-G) Data shown as average ± standard error of the mean (SEM) for TILS (red) and sham (gray) conditions. Effect sizes and p-values from mixed-effects analyses. *p<0.05, **p<0.01, ***p<0.001, ****p<0.0001.

Analysis of physiological measurements revealed significant temporal changes in HR, VO_2_, and mean arterial pressure (MAP) throughout the cognitive stress experiments (effect of time p<0.0001 for all variables). We did not observe a difference between TILS and sham conditions (Figure 2B-D). HR and MAP demonstrated distinct response patterns, with peaks occurring at the initiation of the MSIT (blood pressure was not measured during the conditioning phase). HR returned to baseline levels shortly after the initiation of the test, while MAP remained elevated 2-7% above baseline throughout the remainder of the task and recovery periods. VO_2_ showed a gradual increase during baseline and conditioning periods, stabilizing at approximately 12% above baseline during both MSIT and recovery phases. Heart rate variability (as RMSSD) did not change significantly over time and was not different between conditions (Figure 2E). Physiological measurement trajectories were not significantly different between initial and follow-up experiments, although average HR, VO_2_, and MAP tended to be lower on follow-up visits (Figure S3A-C).

Cf-mtDNA exhibited significant temporal changes during the cognitive stress experiments (effect of time p<0.0001; Figure 2F). Trajectories were not different between TILS and sham treatment, or between initial and follow-up visits (Figure 2F, Figure S3E). On average, a continuous increase was observed between the initial baseline time point and the final conditioning time point, peaking at +166% above baseline. This was followed by a rapid decrease to 39% above baseline at MSIT initiation and then a secondary increase to 160% above baseline during the task. Finally, we observed a gradual decline during the recovery period, stabilizing within 6-31% above baseline values.

As in the exercise experiment, cf-nDNA varied significantly over time in cognitive stress experiments (p<0.0001) and followed a trajectory similar to that of cf-mtDNA (Figure 2G). Cf-nDNA trajectories were not significantly different between condition (TILS vs. sham) (Figure 2G) or visit order (Figure S3F).

When physiological and cf-DNA measurements from the cognitive stress experiment were normalized to baseline values, all variables except RMSSD varied significantly over time and were not significantly different between TILS and sham conditions (Figure S4).

The cf-mtDNA/cf-nDNA ratio changed significantly over the cognitive stress experiment (p=0.0014; Figure S2C), decreasing between baseline and the middle of the stress task and then gradually increasing towards baseline during the recovery period. When cf-mtDNA/cf-nDNA ratios were normalized to baseline values, they exhibited a similar trajectory and changed significantly over time (p=0.0033) (Figure S2D).

### 3.3. Determinants of saliva cf-mtDNA responses

Cf-mtDNA trajectories varied dramatically between participants in both the exercise and cognitive stress experiments. To explore factors that may shape cf-mtDNA responses and identify potential subgroups with similar cf-mtDNA trajectory patterns, trajectories from exercise experiments, initial cognitive stress experiments, and follow-up cognitive stress experiments were separately fitted in LCLMM. Trajectories from TILS and sham conditions were combined because trajectories did not differ significantly between conditions (see Figure 2F). For each experiment, cf-mtDNA trajectories with 10 or more measurements were assigned to one of two latent classes (Table 2). Before clustering, cf-mtDNA measurements in each individual trajectory were normalized by dividing by their average, thereby allowing trajectories to be grouped by their relative shape instead of absolute values. To elucidate potential determinants of the observed response trajectories, we conducted a comprehensive analysis of demographic and behavioral characteristics across participant groups, revealing several statistically significant differences between classes (Table 3).

**Table 2.** Participant class assignments from latent class linear mixed models for each experiment (exercise, initial cognitive stress experiment, follow-up cognitive stress experiment)

| LCLMM Class Assignments |  |  |  |
| --- | --- | --- | --- |
| Participant ID | Exercise experiment | Initial Cognitive stress experiment | Follow-Up Cognitive stress experiment |
| 3 | 2 | 2 | 2 |
| 4 | 2 | (No experiment) | (No experiment) |
| 5 | 1 | 2 | 1 |
| 6 | 1 | 2 | 2 |
| 7 | 2 | 2 | 2 |
| 8 | 2 | 2 | 2 |
| 9 | 2 | (No experiment) | (No experiment) |
| 10 | 2 | 2 | 2 |
| 11 | (Too few samples for modeling) | (Samples not sent) | 1 |
| 12 | 2 | 1 | (No experiment) |
| 13 | 2 | (No experiment) | (No experiment) |
| 14 | (Samples not sent) | 1 | (Samples not sent) |
| 15 | 1 | 2 | 1 |
| 16 | (Too few samples for modeling) | 2 | (Too few samples for modeling) |
| 17 | (Too few samples for modeling) | 2 | 1 |
| 18 | 2 | 2 | 1 |
| 19 | 1 | 1 | 1 |
| 24 | 2 | 2 | 1 |
| 25 | 1 | (No experiment) | (No experiment) |
| 26 | 2 | 2 | 1 |
| 27 | 2 | 2 | 1 |
| 28 | (Too few samples for modeling) | 2 | 2 |
| 29 | 2 | 1 | 1 |
| 30 | 2 | 1 | 1 |
| 31 | (Too few samples for modeling) | 1 | 1 |
| 32 | (Too few samples for modeling) | 1 | 2 |
| 33 | 1 | 1 | 2 |
| 35 | 2 | 1 | 1 |
| 36 | 2 | 2 | 1 |
| 37 | 1 | 1 | 1 |
| 38 | 2 | 1 | 2 |
| 39 | 2 | 2 | (Samples not sent) |
| 40 | 1 | (No experiment) | (No experiment) |
| 41 | (Samples not sent) | (Samples not sent) | 1 |
| 42 | (Too few samples for modeling) | (Too few samples for modeling) | 2 |
| 43 | (Too few samples for modeling) | 2 | 1 |
| 44 | 2 | 1 | 2 |
| 46 | 2 | 2 | 2 |
| 47 | 2 | (No experiment) | (No experiment) |
| 48 | 2 | 1 | 1 |
| 61 | 1 | 2 | 2 |
| 62 | (Too few samples for modeling) | 2 | 2 |
| 63 | (Too few samples for modeling) | 2 | 1 |
| 64 | 2 | 2 | (Samples not sent) |
| 65 | (Too few samples for modeling) | (Too few samples for modeling) | (Too few samples for modeling) |
| 66 | (Too few samples for modeling) | (Too few samples for modeling) | (Too few samples for modeling) |

**Table 3.** P values and effect sizes from comparisons of biometric and psychosocial variables between participants assigned to different latent classes based on their cf-mtDNA trajectories, separated by experiment.

| Variable | Exercise Experiment |  | Initial Cognitive Stress Experiment |  | Follow-Up Cognitive Stress Experiment |  |
| --- | --- | --- | --- | --- | --- | --- |
|  | *p*-value | Effect size<br>(Class 1 vs.<br>Class 2) | *p*-value | Effect size<br>(Class 1 vs.<br>Class 2) | *p*-value | Effect size<br>(Class 1 vs.<br>Class 2) |
| Sex<sup>†</sup> | 0.70 | 0.00 | 0.49 | 0.00 | 0.73 | 0.00 |
| Age | 0.15 | -0.45 | 0.88 | 0.05 | 0.36 | 0.29 |
| BMI | 0.01* | -0.71 | 0.93 | -0.03 | 0.76 | -0.11 |
| Mental health condition (yes / no)<sup>†</sup> | 1.00 | 0.00 | 0.13 | 0.22 | 0.63 | 0.00 |
| Medication (yes / no)<sup>†</sup> | 0.15 | 0.24 | 0.68 | 0.00 | 0.12 | 0.24 |
| Birth control (yes / no)<sup>†</sup> | 0.08 | 0.46 | 0.62 | 0.00 | 1.00 | 0.00 |
| Cigarette smoking frequency<sup>‡</sup> | 0.53 | 0.11 | 0.19 | 0.22 | 0.38 | 0.15 |
| Cannabis smoking frequency<sup>‡</sup> | 0.81 | 0.36 | 0.09 | 0.30 | 0.74 | 0.06 |
| Alcohol use frequency<sup>‡</sup> | 0.03* | 0.53 | 0.30 | 0.35 | 0.84 | 0.49 |
| Physical activity level<sup>‡</sup> | 0.86 | 0.32 | 0.55 | 0.33 | 0.05 | 0.44 |
| PSQI score (total) | 0.94 | -0.03 | 0.17 | -0.49 | 0.03* | 0.82 |
| HADS anxiety | 0.49 | -0.31 | 0.13 | -0.59 | 0.86 | -0.06 |
| HADS depression | 0.77 | 0.13 | 0.59 | -0.21 | 0.47 | -0.28 |
| Perceived stress scale | 0.35 | -0.48 | 0.58 | -0.21 | 0.87 | -0.06 |
| ERQ reappraisal | 0.17 | -0.65 | 0.23 | 0.49 | 0.39 | -0.30 |
| ERQ suppression | 0.77 | -0.13 | 0.54 | -0.23 | 0.47 | -0.24 |
| Adverse childhood experiences | 0.09 | 0.77 | 0.38 | -0.36 | 0.42 | 0.30 |
| USQ total | 0.02* | -1.16 | 0.71 | -0.15 | 0.40 | 0.29 |
| USQ total (weighted) | 0.02* | -1.16 | 0.75 | -0.13 | 0.40 | 0.29 |
| Exercise max VO<sub>2</sub> | 0.16 | 0.57 | 0.79 | 0.08 | 0.88 | 0.05 |
| Pulse wave velocity (Visit 1) | 0.04* | -0.78 | 0.77 | -0.12 | 0.09 | 0.61 |
| SBP reactivity<sup>§</sup> | *n.a.* | *n.a.* | 0.89 | -0.05 | 0.07 | 0.67 |
| DBP reactivity<sup>§</sup> | *n.a.* | *n.a.* | 0.62 | 0.16 | 0.01** | 0.95 |
| MAP reactivity<sup>§</sup> | *n.a.* | *n.a.* | 0.98 | -0.01 | 0.01** | 0.92 |
| VO<sub>2</sub> reactivity<sup>§</sup> | 0.06 | 0.78 | 0.27 | -0.41 | 0.05* | 0.69 |
| Heart rate reactivity<sup>§</sup> | 0.85 | -0.08 | 0.44 | -0.25 | 0.09 | 0.53 |
| Heart rate variability reactivity<sup>§</sup> | *n.a.* | *n.a.* | 0.57 | -0.12 | 0.27 | 0.36 |
| 15W exercise VO<sub>2</sub> | 0.21 | 0.69 | 0.97 | -0.01 | 0.54 | 0.19 |
| 30W exercise VO<sub>2</sub> | 0.12 | 0.71 | 0.86 | -0.05 | 0.38 | 0.30 |
| 60W exercise VO<sub>2</sub> | 0.07 | 0.67 | 0.53 | -0.20 | 0.31 | 0.37 |
| 90W exercise VO<sub>2</sub> | 0.04* | 0.75 | 0.57 | -0.19 | 0.50 | 0.24 |

† *p*-value from Fisher's exact test; effect size: Phi; ‡ *p*-value from Armitage trend test; effect size: Cohen's *W*; § reactivity defined as difference between the maximum post-baseline value and the average of baseline time points; *n.a.* blood pressure and heart rate variability not measured during exercise; USQ Undergraduate stress questionnaire; PSQI Pittsburgh sleep quality index; ERQ Emotional response questionnaire; HADS Hospital anxiety and depression scale; \* *p* < 0.05; \*\* *p* < 0.01

#### 3.3.1. Determinants of saliva cf-mtDNA responses to acute exercise stress

In the exercise experiment, Class 1 cf-mtDNA trajectories (n=9) showed a progressive increase throughout the entire experiment, peaking at ∼1.4-fold above baseline at the final time point, while Class 2 trajectories (n=23) exhibited a modest rise (∼1.13-fold) that coincided with the exercise phase, followed by a return to baseline during recovery (Figure 3A). Compared to Class 2 participants, Class 1 participants consumed alcohol less often (only on special occasions or abstaining entirely; 100% vs. 43%, Cohen’s w=0.53, p=0.029), were less likely to be using birth control (female participants only; 0% vs. 38.5%, *phi*=0.46, p=0.082) (Figure 3B), had lower BMI (Hedge’s g=-0.71, p=0.01) (Figure 3C), reported fewer recent stressful events (Undergraduate Stress Questionnaire score; Hedge’s g=-1.16, p=0.02) (Figure 3D), had more total adverse childhood experiences (Hedge’s g=0.77, p=0.09) (Figure 3E), higher VO_2_ reactivity (Hedge’s g=0.78, p=0.06) (Figure 3F), and higher maximum VO_2_ at 90W exercise (Hedge’s g=0.75, p=0.035) (Figure 3G). These findings highlight the potential influence of hormonal factors, alcohol consumption, stress levels and physiological fitness on cf-mtDNA dynamics during exercise stress.

**Figure 3.**
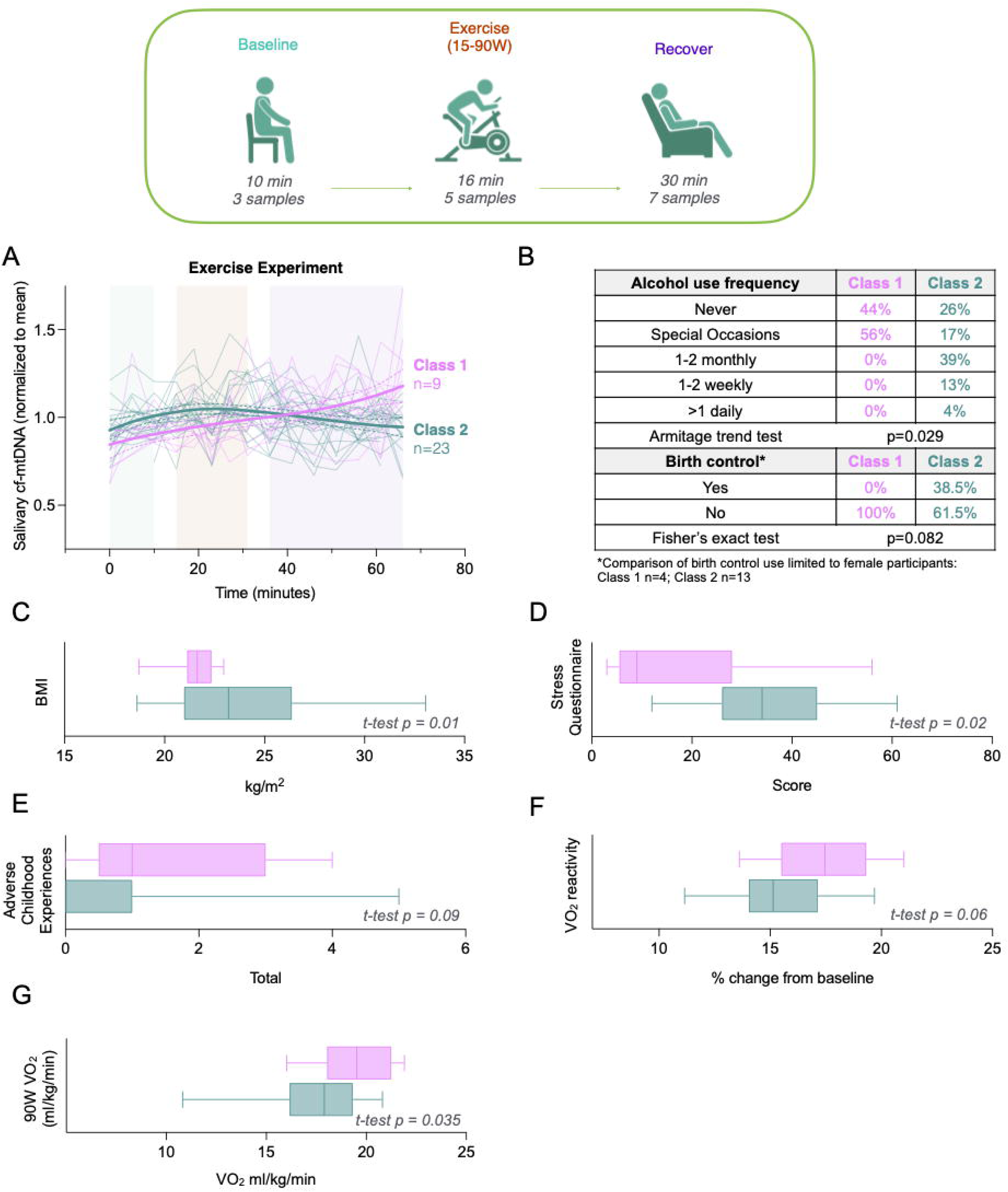
Determinants saliva cf-mtDNA responses to exercise stress. Participants’ mean-normalized trajectories of log-transformed cf-mtDNA abundance over the exercise experiment were grouped into two classes of responses using a latent class linear mixed model. **(A)** Trajectories data shown as the plot of the model determined for each latent class (thick solid line) with 95% confidence interval (dashed lines) and individual mean-normalized trajectories (thin transparent lines). (**B-G)** Characteristics distinguishing the two groups below a statistical threshold of p < 0.1. Box plots (C-G) represent range, interquartile range, and median of values for each class. (D) “Stress Questionnaire” score is from Undergraduate Stress Questionnaire (unweighted). (F) VO_2_ reactivity was calculated as the percent change between baseline and the highest value post-baseline. Effect sizes from (B) Fisher’s exact test, Armitage trend test and (C-G) Welch’s t-test.

#### 3.3.2. Determinants of saliva cf-mtDNA responses to acute cognitive stress

In the initial cognitive stress experiment (first visit, TILS or sham independent) Class 1 trajectories (n=13) had a relatively dynamic saliva cf-mtDNA response while Class 2 trajectories (n=22) were characterized by a flatter response (Figure 4A). No characteristics were significantly different between the groups, although Class 2 participants (flatter trajectories) were more likely to abstain from cannabis use (95% vs. 77%, Cohen’s w=0.30, p=0.09) and reported fewer mental health disorders (4% vs. 25%, *phi*=0.22, p=0.13) (Figure 4B). These findings suggest that differences in participants’ cf-mtDNA dynamics in response to acute cognitive stress could be related to lifestyle factors, such as substance use.

**Figure 4.**
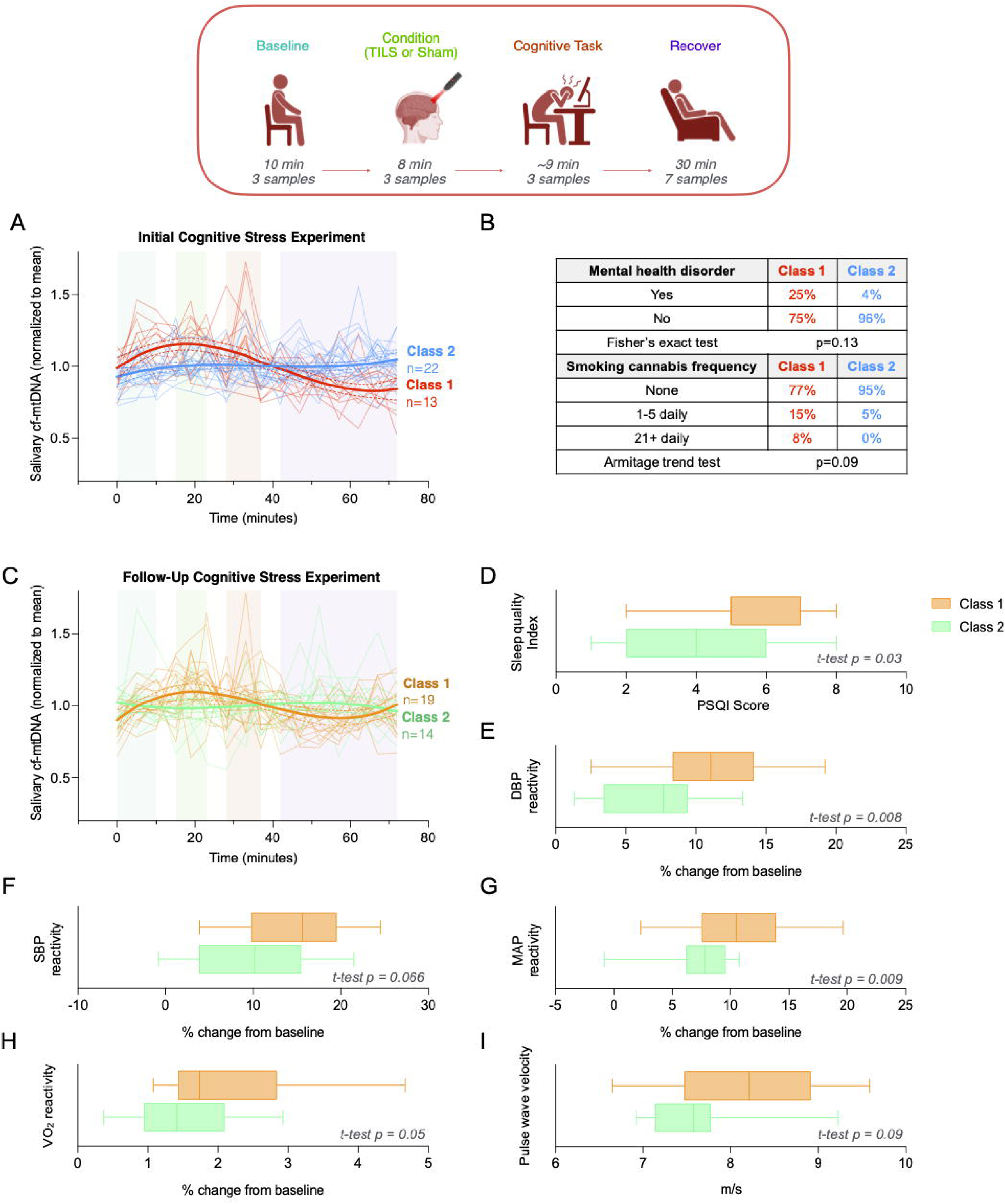
Determinants of saliva cf-mtDNA responses to cognitive stress. **(A)** Participants’ mean-normalized trajectories of log-transformed cf-mtDNA abundance over the initial cognitive stress experiments were grouped into two classes of responses using a latent class linear mixed model. Trajectories data shown as the plot of the model determined for each latent class (thick solid line) with 95% confidence interval (dashed lines) and individual mean-normalized trajectories (thin transparent lines). **(B)** Characteristics distinguishing the two groups. **(C,D-I)** Same as (A,B) for follow-up cognitive stress experiments. Box plots (D-I) represent range, interquartile range, and median of values for each class. “Sleep Quality” score (D) is from Pittsburgh Sleep Quality Index. Blood pressure reactivity (SBP, DBP, MAP) and VO_2_ reactivity (E-H) were calculated as the percent change between baseline and the highest value measured post-baseline. Pulse wave velocity (I) was measured during visit 1. Effect sizes and p-values from (B) Fisher’s exact test or Armitage trend test and (D-I) Welch’s t-test.

Cf-mtDNA trajectories from follow-up cognitive stress experiments were similarly clustered. Class 1 cf-mtDNA trajectories were again characterized as having a more dynamic response (n=19), while Class 2 trajectories had a flatter response (n=14) (Figure 4C). The factors that were most significantly different between participants with Class 1 and Class 2 cf-mtDNA trajectories in the follow-up cognitive stress experiment were not the same as those that differed between classes in the initial cognitive stress experiment. Participants with Class 2 (flatter) trajectories reported better sleep quality (lower Pittsburgh Sleep Quality Index; Hedge’s g=0.82, p=0.03) (Figure 4D) and exhibited lower blood pressure reactivity (systolic, Hedge’s g=0.67, p=0.07; diastolic, Hedge’s g=0.95, p=0.01; mean arterial pressure, Hedge’s g=0.92, p=0.01) (Figure 4E-G), lower VO_2_ reactivity (Hedge’s g=0.69, p=0.05) (Figure 4H), and had lower arterial stiffness (pulse wave velocity; Hedge’s g=0.63, p=0.09) (Figure 4I) suggesting that physiological reactivity and sleep quality may play a role in shaping cf-mtDNA responses to acute cognitive stress.

## Discussion

This study provides novel insights into saliva cf-mtDNA responses to acute physical and cognitive stress. First, we showed that saliva cf-mtDNA increases after both stress paradigms, reflecting dynamic mitochondrial signaling across stress modalities. Second, we identified distinct cf-mtDNA response trajectories between subsets of participants and performed an exploratory analysis to study their potential behavioral, physiological, and demographic determinants. Participants with lower BMI, lower reported stress, and greater stressor-evoked VO_2_ responses showed larger cf-mtDNA responses to exercise, whereas greater reactivity to cognitive stress was linked to poorer sleep quality and greater stressor-evoked cardiovascular responses. These findings contribute to the growing body of evidence supporting saliva cf-mtDNA as a promising stress biomarker^17,19,20,36,37^.

During the exercise protocol, cf-mtDNA levels varied significantly, increasing by 1.9-fold on average immediately following baseline and remained elevated throughout the experiment, independent of exercise intensity. Previous studies investigating the effects of exercise stress on blood cf-mtDNA have yielded mixed results. Some demonstrate an increase in plasma cf-mtDNA levels following acute exercise stress^14,16^, while others show a decrease^38^ (reviewed in Trumpff et al. 2021^12^). A recent study found that saliva cf-mtDNA levels were not affected by acute exercise stress^17^. Overall, our results suggest that saliva cf-mtDNA levels show a modest increase following exercise stress, an effect that remains to be confirmed in future studies.

In the cognitive stress experiments, cf-mtDNA levels showed dynamic fluctuations, with peaks observed during the conditioning and task phases. On average, cf-mtDNA increased approximately 2.6-fold between baseline (0 minutes) and the middle of the MSIT (33 minutes). These findings are consistent with prior studies which found cf-mtDNA to increase following acute psychological stress in blood^15,16,18,20^ and saliva^19,20^. One study demonstrated plasma cf-mtDNA to increase by approximately 1.3-fold following a socio-evaluative stressor^20^, while another demonstrated a 1.6-fold increase in plasma cf-mtDNA following a similar stressor^17^. These same two studies also demonstrated 10-fold^20^ and 1.5-fold^17^ increases in saliva cf-mtDNA following the same stimuli. However, a recent study demonstrated similar reactivities of cf-mtDNA in plasma following both a socio-evaluative stressor and a control condition where participants sat quietly, calling into question whether other aspects of the study were more important than the stressor in stimulating cf-mtDNA dynamics^18^. The differences between our results and those of other studies investigating saliva cf-mtDNA responses to acute psychological stress may be attributable to the stressor used^17,20^ and sample preparation methods^17^. The socio-evaluative stressor used in Trumpff et al. 2025^18^ elicited a 1.33-fold increase in heart rate above baseline, while the MSIT only increased heart rate by 1.13-fold. The sample preparation method used in Limberg et al. (2025)^17^ involved a lysis buffer that did not include a detergent, which may have resulted in a different population of cf-mtDNA being measured than in our study.

We found substantial inter-individual variability in cf-mtDNA trajectories in both exercise and cognitive stress experiments. Using LCLMM, we categorized trajectories as one of two classes in each experiment. Participants with sustained cf-mtDNA elevations during exercise were characterized by lower BMI, less frequent alcohol use, higher VO_2_ max during intense exercise, and lower perceived stress levels. These findings suggest that fitness and stress resilience may be associated with larger saliva cf-mtDNA release during exercise stress. In initial cognitive stress visits, no factors were significantly different between participants with more dynamic and ‘flatter’ cf-mtDNA trajectories, although those with larger responses were somewhat more likely to use cannabis or have a mental health condition. During follow-up cognitive stress experiments, participants with larger cf-mtDNA responses were more likely to report poorer sleep quality and exhibited greater blood pressure reactivity, highlighting the potential influence of physiological and behavioral factors on stress reactivity. The observed differences in cf-mtDNA trajectories between the initial and follow-up cognitive stress experiments further underscore the dynamic nature of stress responses. While lifestyle factors such as substance use were associated with larger cf-mtDNA responses during the initial visit, greater physiological reactivity and poorer sleep quality emerged as significant predictors of a more dynamic response during the follow-up session. These findings suggest that the determinants of cf-mtDNA responses may vary depending on the context and timing of the stressor, reflecting the complex and multifaceted nature of stress adaptation.

The identification of saliva cf-mtDNA as a dynamic and context-sensitive biomarker of stress has important implications for both research and clinical practice. First, the non-invasive nature of saliva cf-mtDNA measurement makes it a practical and accessible tool for monitoring stress responses in real-world settings. The sensitivity and relative affordability of qPCR also present an advantage over other analytical techniques. On the other hand, the relationship between saliva cf-mtDNA dynamics and other stress responses is not well understood, nor is the physiological significance of the saliva cf-mtDNA response. Our results suggest that more physically fit individuals are capable of mounting a gradual but overall larger saliva cf-mtDNA response to exercise stress, suggesting this phenomenon might be adaptative.

Although cell-free mitochondrial and nuclear DNA trajectories exhibited a small degree of independence, as can be observed from the changes in their ratio over the exercise and cognitive stress experiments (Figure S2), they largely matched one another. This is consistent with our previous observations of cf-DNA in saliva^19,20,36^. This contrasts with observations in plasma and serum, where cf-mtDNA and cf-nDNA trajectories in response to acute psychosocial stress are weakly correlated^15,18,20^. In blood, an unidentified mechanism involving the active secretion of membrane-encapsulated mtDNA, or whole mitochondria, has been hypothesized as the source of cf-mtDNA^8,12,39^. The similar trajectories of cf-mtDNA and cf-nDNA in saliva over experimental time courses suggests that a mechanism different from that in blood drives their dynamics. The oral epithelium is in a constant state of shedding and renewal^40^. The lysis of these epithelial cells could explain the apparent co-regulation of cf-mtDNA and cf-nDNA in saliva. However, because the levels of both types of saliva cf-DNA change significantly over time in both exercise and stress experiments, they still hold potential as valuable biomarkers of acute physical and psychological or cognitive stress. More research is necessary to elucidate the sources of saliva cf-DNA and how they differ from those of serum and plasma.

The potential buffering effects of transcranial infrared laser stimulation (TILS) on stress responses warrant further investigation. Although we did not observe significant differences in cf-mtDNA trajectories between TILS and sham conditions, this remains to be confirmed by other studies with different experimental stressors and with larger sample sizes. Coupling TILS conditioning with socio-evaluative stress experiments^17,18,20^, rather than cognitive stress experiments, could be a worthwhile direction for future studies. We have previously demonstrated that acute socio-evaluative stress can elicit an increase in cf-mtDNA in saliva and blood^18,20^. Given that TILS is being explored as a therapeutic option for anxiety and mood disorders^41,42^, it could be worthwhile to study whether TILS either pre- or post-psychosocial stress modulates physiological responses, including cf-mtDNA dynamics.

While this study provides valuable insights into cf-mtDNA responses to acute stress, several limitations should be acknowledged. First, the sample size, though sufficient for detecting significant effects, may limit the generalizability of our findings. Larger, more diverse studies are needed to validate these results and explore potential sex- and age-related differences in cf-mtDNA responses. Second, the observational nature of this study precludes causal inferences about the relationships between individual characteristics and cf-mtDNA trajectories. Experimental studies manipulating key determinants, such as fitness or sleep quality, are needed to establish causality. Third, while we focused on saliva cf-mtDNA, future research should investigate its relationship with circulating cf-mtDNA and other biomarkers of stress to provide a more comprehensive understanding of stress physiology. Finally, the biological mechanisms underlying cf-mtDNA release during stress remain poorly understood. While our findings suggest that cf-mtDNA responses are influenced by both physiological and behavioral factors, the specific cellular processes driving its release require further investigation. Elucidating these mechanisms will be critical for understanding the role of cf-mtDNA in stress-related health outcomes.

In summary, this study highlights the potential of saliva cf-mtDNA as a biomarker of acute physiological and cognitive stress and underscores the importance of inter-individual differences in shaping stress responses.

## Acknowledgements

This study was supported by NIH K01HL145021 to A.G. The study also benefited from support from the Columbia Irving Institute Scholars Program, the Wharton Fund, and the Baszucki Group to M.P.

## Data availability

Data will be made available upon request.

## Author contributions

A.G. designed the original study and secured funding. M.P. and C.T. added cf-mtDNA to the study design. A.G. oversaw all data collection. D.S. and T.Y. performed the cf-mtDNA assays. D.S., T.W., and S.W. ran the statistical analysis. C.T. and D.S. wrote the manuscript. All authors reviewed the final version of the manuscript.

## References

1 McEwen, B. S. Stress, adaptation, and disease: Allostasis and allostatic load. Annals of the New York Academy of Sciences 840, 33–44 (1998).

2 Cohen, S., Janicki-Deverts, D. & Miller, G. E. Psychological stress and disease. Jama 298, 1685–1687 (2007).

3 Kelly, C. et al. A platform to map the mind–mitochondria connection and the hallmarks of psychobiology: the MiSBIE study. Trends in Endocrinology & Metabolism 35, 884–901 (2024).

4 Picard, M., McEwen, B. S., Epel, E. & Sandi, C. An energetic view of stress: Focus on mitochondria. Frontiers in neuroendocrinology (2018).

5 Miliotis, S., Nicolalde, B., Ortega, M., Yepez, J. & Caicedo, A. Forms of extracellular mitochondria and their impact in health. Mitochondrion 48, 16–30 (2019).

6 Brestoff, J. R. et al. Recommendations for mitochondria transfer and transplantation nomenclature and characterization. Nature Metabolism, 1–15 (2025).

7 Bobba-Alves, N., Juster, R.-P. & Picard, M. The energetic cost of allostasis and allostatic load. Psychoneuroendocrinology 146, 105951 (2022).

8 Al Amir Dache, Z., et al. Blood contains circulating cell-free respiratory competent mitochondria. The FASEB Journal, 3616–3630 (2020).

9 D’Acunzo, P. et al. Mitovesicles are a novel population of extracellular vesicles of mitochondrial origin altered in Down syndrome. Science advances 7, eabe5085 (2021).

10 Song, X. et al. Existence of circulating mitochondria in human and animal peripheral blood. International Journal of Molecular Sciences 21, 2122 (2020).

11 Caicedo, A. et al. Decoding the nature and complexity of extracellular mtDNA: Types and implications for health and disease. Mitochondrion 75, 101848 (2024).

12 Trumpff, C. et al. Stress and circulating cell-free mitochondrial DNA: A systematic review of human studies, physiological considerations, and technical recommendations. Mitochondrion 59, 225–245 (2021).

13 Stawski, R. et al. Repeated bouts of exhaustive exercise increase circulating cell free nuclear and mitochondrial DNA without development of tolerance in healthy men. PloS one 12, e0178216 (2017).

14 Ohlsson, L. et al. Increased level of circulating cell-free mitochondrial DNA due to a single bout of strenuous physical exercise. European Journal of Applied Physiology 120, 897–905 (2020).

15 Trumpff, C. et al. Acute psychological stress increases serum circulating cell-free mitochondrial DNA. Psychoneuroendocrinology 106, 268–276 (2019).

16 Hummel, E. et al. Cell-free DNA release under psychosocial and physical stress conditions. Translational Psychiatry 8, 236 (2018).

17 Limberg, A. et al. Cell-free DNA release following psychosocial and physical stress in women and men. Translational Psychiatry 15, 26 (2025).

18 Trumpff, C. et al. Effects of acute psychological stress on blood cell-free mitochondrial DNA (cf-mtDNA): A crossover experimental study. Psychoneuroendocrinology 182, 107644 (2025). 10.1016/j.psyneuen.2025.107644

19 Trumpff, C. et al. Dynamic behavior of cell-free mitochondrial DNA in human saliva. Psychoneuroendocrinology 143, 105852 (2022).

20 Trumpff, C. et al. Saliva and blood cell-free mtDNA reactivity to acute psychosocial stress. Psychoneuroendocrinology 179, 107506 (2025). 10.1016/j.psyneuen.2025.107506

21 Salehpour, F. et al. Brain photobiomodulation therapy: a narrative review. Molecular neurobiology 55, 6601–6636 (2018).

22 Zaizar, E. D., Gonzalez-Lima, F. & Telch, M. J. Singular and combined effects of transcranial infrared laser stimulation and exposure therapy: A randomized clinical trial. Contemp Clin Trials 72, 95–102 (2018). 10.1016/j.cct.2018.07.012

23 Buysse, D. J., Reynolds III, C. F., Monk, T. H., Berman, S. R. & Kupfer, D. J. The Pittsburgh Sleep Quality Index: a new instrument for psychiatric practice and research. Psychiatry research 28, 193–213 (1989).

24 Zigmond, A. S. & Snaith, R. P. The hospital anxiety and depression scale. Acta psychiatrica scandinavica 67, 361–370 (1983).

25 Cohen, S., Kamarck, T. & Mermelstein, R. A global measure of perceived stress. Journal of health and social behavior, 385–396 (1983).

26 Gross, J. J. & John, O. P. Individual differences in two emotion regulation processes: implications for affect, relationships, and well-being. Journal of personality and social psychology 85, 348 (2003).

27 Anda, R. F. et al. The enduring effects of abuse and related adverse experiences in childhood: A convergence of evidence from neurobiology and epidemiology. European archives of psychiatry and clinical neuroscience 256, 174–186 (2006).

28 Crandall, C. S., Preisler, J. J. & Aussprung, J. Measuring life event stress in the lives of college students: The Undergraduate Stress Questionnaire (USQ). Journal of behavioral medicine 15, 627–662 (1992).

29 Bush, G., Shin, L., Holmes, J., Rosen, B. & Vogt, B. The Multi-Source Interference Task: validation study with fMRI in individual subjects. Molecular psychiatry 8, 60–70 (2003).

30 Sheu, L. K., Jennings, J. R. & Gianaros, P. J. Test–retest reliability of an fMRI paradigm for studies of cardiovascular reactivity. Psychophysiology 49, 873–884 (2012).

31 Ginty, A. T., Gianaros, P. J., Derbyshire, S. W., Phillips, A. C. & Carroll, D. Blunted cardiac stress reactivity relates to neural hypoactivation. Psychophysiology 50, 219–229 (2013).

32 Tyra, A. T., Garner, S.-B. & Ginty, A. T. Examining the association between habitual emotion regulation strategies and cardiovascular stress reactivity across three studies. Biological psychology 194, 108966 (2025).

33 Gianaros, P. J. et al. Effects of a year-long aerobic exercise intervention on neuroendocrine, autonomic, and neural correlates of stress, emotion, and cardiovascular disease risk in midlife adults. Journal of Sport and Health Science, 101135 (2026).

34 Michelson, J. et al. MitoQuicLy: A high-throughput method for quantifying cell-free DNA from human plasma, serum, and saliva. Mitochondrion 71, 26–39 (2023). 10.1016/j.mito.2023.05.001

35 Burnham, K. P. & Anderson, D. R. in Model selection and inference: a practical information-theoretic approach 32–74 (Springer, 1998).

36 Behnke, A. et al. Diurnal dynamics and psychobiological regulation of cell-free mitochondrial and nuclear DNA in human saliva. medRxiv, 2025.2010. 2013.25337887 (2025).

37 Behnke, A. et al. Diurnal dynamics and psychobiological regulation of cell-free mitochondrial and nuclear DNA in human saliva. *Brain*, Behavior, and Immunity, 106825 (2026).

38 Shockett, P. E. et al. Plasma cell-free mitochondrial DNA declines in response to prolonged moderate aerobic exercise. Physiological reports 4, e12672 (2016).

39 Stephens, O. R. et al. Characterization and origins of cell-free mitochondria in healthy murine and human blood. Mitochondrion 54, 102–112 (2020).

40 Thomson, P. Back to the future: revisiting oral carcinogenesis, stem cells and epithelial cell proliferation. Faculty Dental Journal 11, 30–34 (2020).

41 Caldieraro, M. A. & Cassano, P. Transcranial and systemic photobiomodulation for major depressive disorder: A systematic review of efficacy, tolerability and biological mechanisms. Journal of affective disorders 243, 262–273 (2019).

42 Cassano, P., Petrie, S. R., Hamblin, M. R., Henderson, T. A. & Iosifescu, D. V. Review of transcranial photobiomodulation for major depressive disorder: targeting brain metabolism, inflammation, oxidative stress, and neurogenesis. Neurophotonics 3, 031404–031404 (2016).

