## Supplemental Figures for "Saliva cell-free mitochondrial DNA (cf-mtDNA) response during physical and cognitive stress"

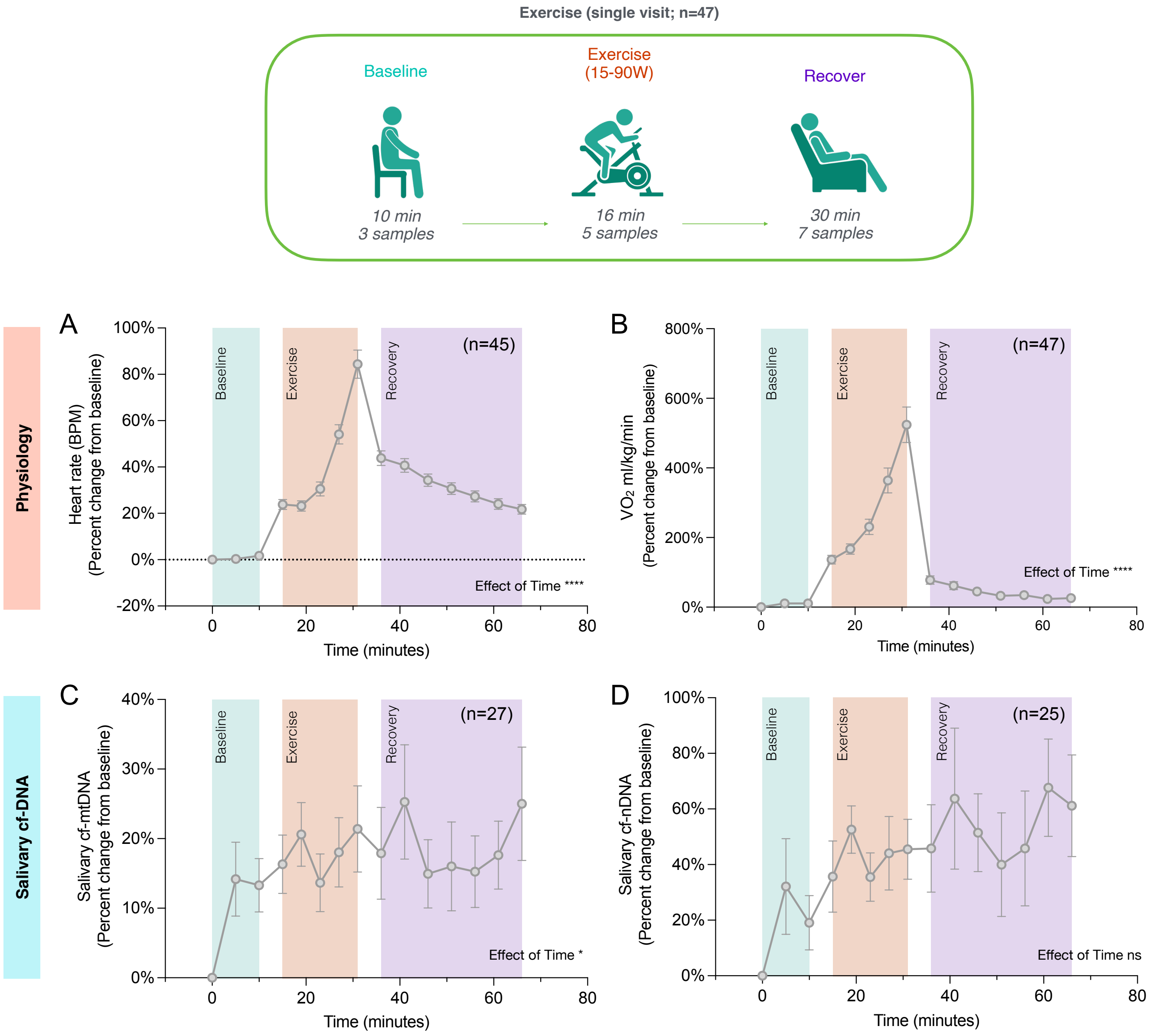


**Supplemental Figure 1. Baseline-normalized trajectories for physiological and saliva cf-DNA responses to exercise stress.** Trajectories of **(A)** heart rate, **(B)** oxygen consumption ($VO_2$ ml/kg/min), **(C)** salivary cf-mtDNA and **(D)** salivary cf-nDNA abundance over exercise visits. Values represent average percent change relative to baseline (t=0) at each time point. Data shown as average ± standard error of the mean (SEM). Effect sizes and p-values from one-way ANOVA. *p<0.05, **p<0.01, ***p<0.001, ****p<0.0001.

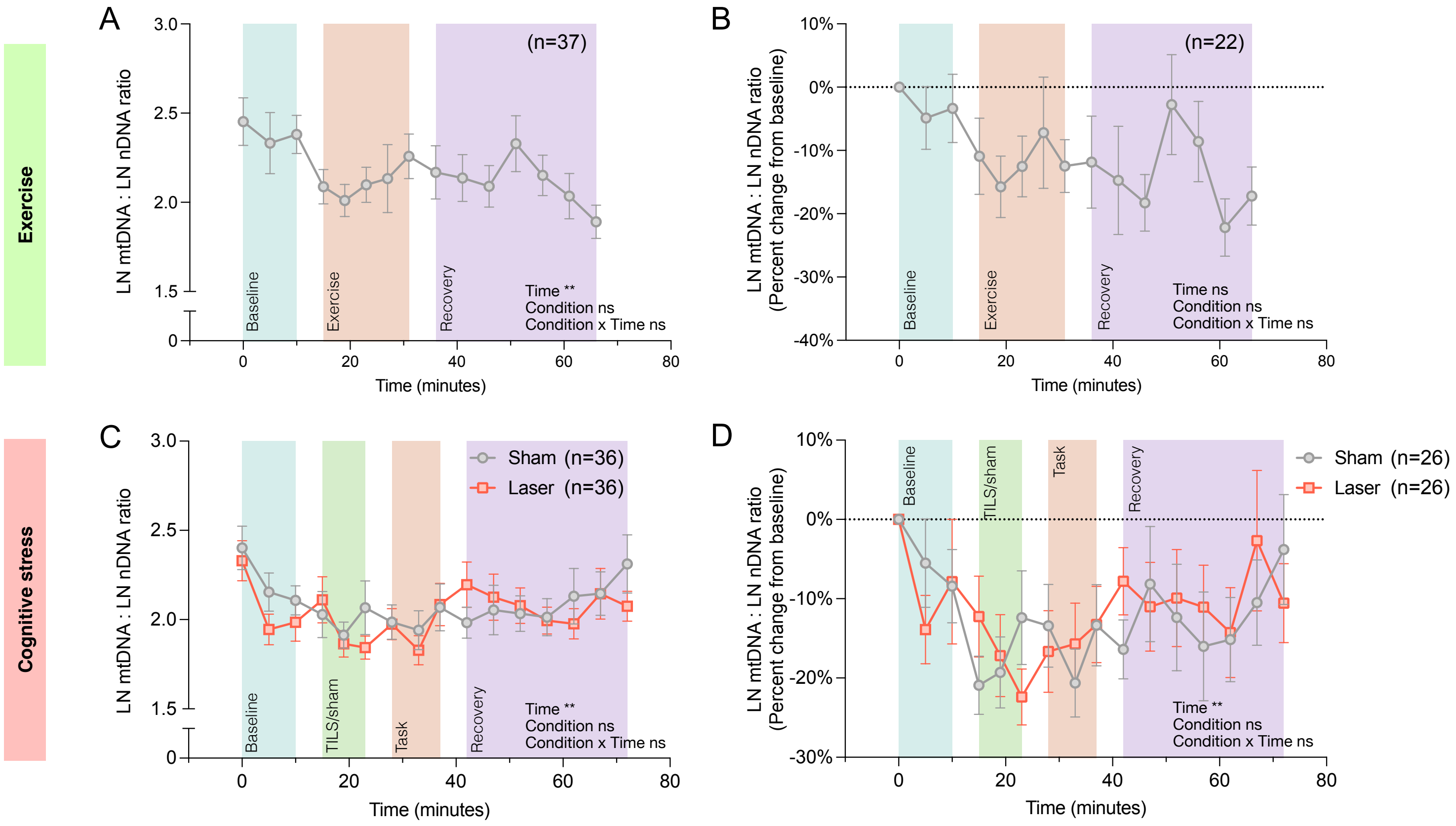


**Supplemental Figure 2. Saliva cf-mtDNA : cf-nDNA ratio trajectories in response to exercise and cognitive stress. (A,B)** Saliva cf-mtDNA to cf-nDNA ratio over the exercise stress experiments. Values represent **(A)** the average ratio of log-transformed cf-mtDNA and cf-nDNA abundance and **(B)** the percent change of this ratio relative to baseline (t=0). **(C,D)** Same as (A,B) for cognitive stress experiments. Trajectories are separated by TILS (red) and sham (gray) conditions. Data shown as average ± standard error of the mean (SEM). Effect sizes and p-values from mixed-effects analyses. *p<0.05, **p<0.01, ***p<0.001, ****p<0.0001.

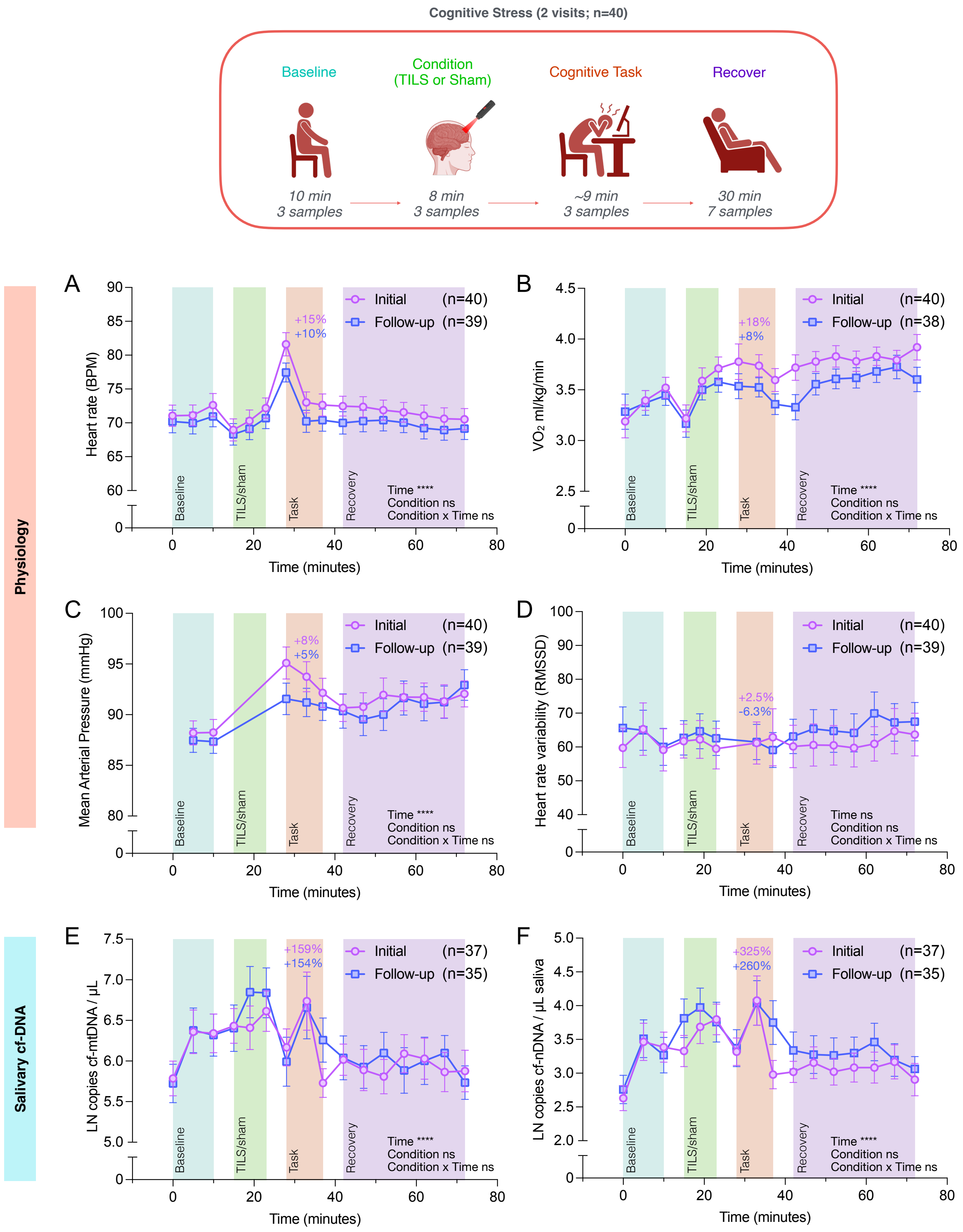


**Supplemental Figure 3. Physiological and cf-DNA trajectories during initial and follow-up cognitive stress experiments.** Trajectories of **(A)** heart rate **(B)** oxygen consumption ($VO_2$ ml/kg/min), **(C)** mean arterial pressure, and **(D)** heart rate variability (as root mean square of successive differences; RMSSD) over the cognitive stress experiments. Values represent averages of several measurements recorded between indicated time points (details are provided in Table S2). Percentages indicate difference between baseline value (t=0) and value at first task time point (t=28). Salivary **(E)** cf-mtDNA and **(F)** cf-nDNA trajectories over the cognitive stress experiments. Percentages indicate difference between geometric mean of baseline values (t=0) and values at second task time point (t=33). (A-F) Data shown as average ± standard error of the mean (SEM) for initial (purple) and follow-up (blue) visits. Effect sizes and p-values from mixed-effects analyses. *p<0.05, **p<0.01, ***p<0.001, ****p<0.0001.

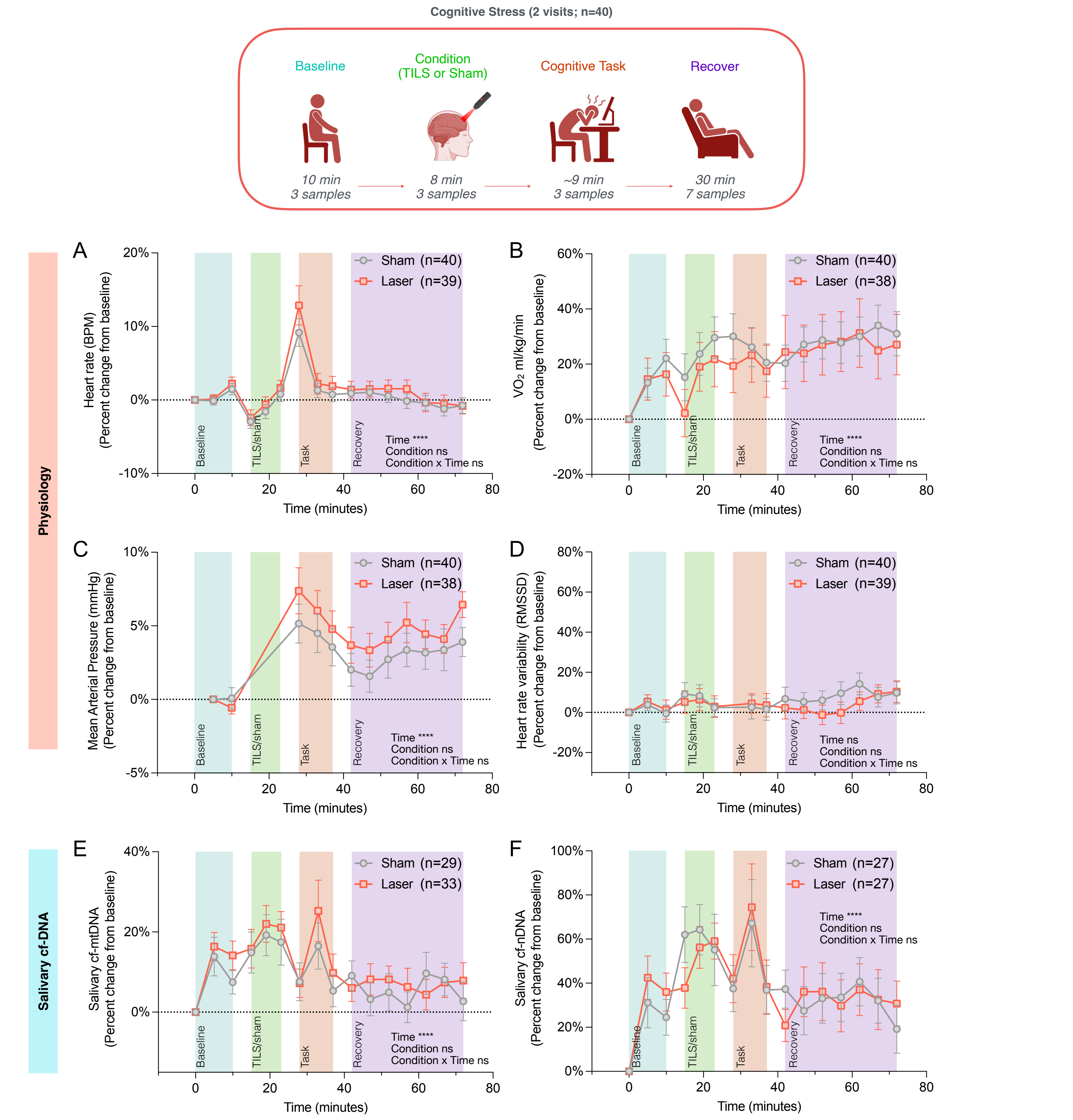


**Supplemental Figure 4. Baseline-normalized trajectories for physiological and saliva cf-DNA responses to cognitive stress.** Trajectories of **(A)** heart rate, **(B)** oxygen consumption ($VO_2$ ml/kg/min), **(C)** mean arterial pressure, **(D)** heart rate variability (as root mean square of successive differences; RMSSD), **(E)** salivary cf-mtDNA, and **(F)** salivary cf-nDNA over the cognitive stress experiments. Values represent average percent change relative to baseline (t=0) at each time point. Data shown as average ± standard error of the mean (SEM) for TILS (red) and sham (gray) conditions. Effect sizes and p-values from mixed-effects analyses. *p<0.05, **p<0.01, ***p<0.001, ****p<0.0001.
