## Supplemental Tables for "Saliva cell-free mitochondrial DNA (cf-mtDNA) response during physical and cognitive stress"

**Table S1. Sequences of qPCR primers and probes**

| **qPCR Target** | **Sequence (5′ → 3′)** | **Concentration in qPCR reaction** |
| --- | --- | --- |
| mt-ND1-F | GAGCGATGGTGAGAGCTAAGGT | 0.3 µM |
| mt-ND1-R | CCCTAAAACCCGCCACATCT | 0.3 µM |
| mt-ND1-Probe | HEX-CCATCACCCTCTACATCACCGCCC-3IABkFQ | 0.15 µM |
| B2M-F | TCTCTCTCCATTCTTCAGTAAGTCAACT | 0.3 µM |
| B2M-R | CCAGCAGAGAATGGAAAGTCAA | 0.3 µM |
| B2M-Probe | FAM-ATGTGTCTGGGTTTCATCCATCCGACA-3IABkFQ | 0.15 µM |

**Table S2. Description of time points and sampling scheme**

Measurements included in each time point for the following variables (measured continuously): Heart Rate, RMSSD, VO_2_ ml/min/kg

**TILS and Sham Visits**

| **Time** | **Phase** | **Time Point/Saliva Sample** | **Measurements** |
| --- | --- | --- | --- |
| 0 min | Baseline | 1 | Min 1 |
| 5 min | Baseline | 2 | Min 1, 2, 3, 4, 5 |
| 10 min | Baseline | 3 | Min 6, 7, 8, 9, 10 |
| 0 min | Laser/Sham | 4 | Min 1 laser |
| 4 min | Laser/Sham | 5 | Min 1, 2, 3, 4 laser |
| 8 min | Laser/Sham | 6 | Min 5, 6, 7, 8 laser |
| 0 min | Stress Task | 7 | Min 1 stress |
| 5 min | Stress Task | 8 | Min 1, 2, 3, 4, 5 stress |
| ~8 min (end of task) | Stress Task | 9 | Min 6, 7, 8 stress |
| 0 min | Recovery | 10 | Min 1 recovery |
| 5 min | Recovery | 11 | Min 1, 2, 3, 4, 5 recovery |
| 10 min | Recovery | 12 | Min 6, 7, 8, 9, 10 recovery |
| 15 min | Recovery | 13 | Min 11, 12, 13, 14, 15 recovery |
| 20 min | Recovery | 14 | Min 16, 17, 18, 19, 20 recovery |
| 25 min | Recovery | 15 | Min 21, 22, 23, 24, 25 recovery |
| 30 min | Recovery | 16 | Min 26, 27, 28, 29, 30 recovery |

Measurements included in each time point for the following variables (measured continuously): SBP, DBP, MAP (taken every 2 minutes during baseline)

**TILS and Sham Visits**

| **Time** | **Phase** | **Time Point/Saliva Sample** | **Measurements** |
| --- | --- | --- | --- |
| 0 min | Baseline | 1 |  |
| 5 min | Baseline | 2 | Min 2, 4 |
| 10 min | Baseline | 3 | Min 6, 8, 10 |
| 0 min | Laser/Sham | 4 |  |
| 4 min | Laser/Sham | 5 |  |
| 8 min | Laser/Sham | 6 |  |
| 0 min | Stress Task | 7 | Min 1 stress |
| 5 min | Stress Task | 8 | Min 2, 3, 4, 5 stress |
| ~8 min (end of task) | Stress Task | 9 | Min 6, 7, 8 stress |
| 0 min | Recovery | 10 | Min 1 recovery |
| 5 min | Recovery | 11 | Min 1, 2, 3, 4, 5 recovery |
| 10 min | Recovery | 12 | Min 6, 7, 8, 9, 10 recovery |
| 15 min | Recovery | 13 | Min 11, 12, 13, 14, 15 recovery |
| 20 min | Recovery | 14 | Min 16, 17, 18, 19, 20 recovery |
| 25 min | Recovery | 15 | Min 21, 22, 23, 24, 25 recovery |
| 30 min | Recovery | 16 | Min 26, 27, 28, 29, 30 recovery |

Measurements included in each time point for the following variables (measured continuously): Heart Rate, RMSSD, VO_2_ ml/min/kg

**Exercise Visit**

| **Time** | **Phase** | **Time Point/Saliva Sample** | **Minutes** |
| --- | --- | --- | --- |
| 0 min | Baseline | 1 | Min 1 |
| 5 min | Baseline | 2 | Min 1, 2, 3, 4, 5 |
| 10 min | Baseline | 3 | Min 6, 7, 8, 9, 10 |
| 0 min | Start Exercise 15W | 4 | Min 1 of 15W |
| 4 min | Start Exercise 30W | 5 | Mins: 1, 2, 3, 4 of 15 W |
| 8 min | Start Exercise 60W | 6 | Min 1, 2, 3, 4 of 30 W |
| 12 min | Start Exercise 90W | 7 | Min 1, 2, 3, 4, of 60 W |
| 16 min | End Required Exercise | 8 | Min 1, 2, 3, 4 of 90 W |
| 0 min | Recovery | 9 | Min 1 recovery |
| 5 min | Recovery | 10 | Min 1, 2, 3, 4, 5 recovery |
| 10 min | Recovery | 11 | Min 6, 7, 8, 9, 10 recovery |
| 15 min | Recovery | 12 | Min 11, 12, 13, 14, 15 recovery |
| 20 min | Recovery | 13 | Min 16, 17, 18, 19, 20 recovery |
| 25 min | Recovery | 14 | Min 21, 22, 23, 24, 25 recovery |
| 30 min | Recovery | 15 | Min 26, 27, 28, 29, 30 recovery |

**Table S3. Goodness-of-fit results and convergence from estimating latent class linear mixed models of cf-mtDNA trajectories using different numbers of latent classes**

| **Experiment** | **Number of classes** | **BIC** | **Model converged?** |
| --- | --- | --- | --- |
| Exercise | 2 | -423.9419 | Yes |
| Exercise | 3 | -378.0629 | No |
| Exercise | 4 | -360.7348 | No |
| Exercise | 5 | -343.4066 | No |
| Cognitive stress (initial) | 2 | -479.6089 | Yes |
| Cognitive stress (initial) | 3 | -461.8224 | No |
| Cognitive stress (initial) | 4 | -444.0483 | No |
| Cognitive stress (initial) | 5 | -426.273 | No |
| Cognitive stress (follow-up) | 2 | -382.4123 | Yes |
| Cognitive stress (follow-up) | 3 | -356.8622 | No |
| Cognitive stress (follow-up) | 4 | -347.7071 | No |
| Cognitive stress (follow-up) | 5 | -321.8986 | No |
